# Intron retention as an excellent marker for diagnosing depression and for discovering new potential pathways for drug intervention

**DOI:** 10.1101/2024.03.30.24305001

**Authors:** Norihiro Okada, Kenshiro Oshima, Akiko Maruko, Mariko Sekine, Naoki Ito, Akino Wakasugi, Eiko Mori, Hiroshi Odaguchi, Yoshinori Kobayashi

## Abstract

**BACKGROUND:** Peripheral inflammation is often associated with depressive disorders, and immunological biomarkers of depression remain a focus of investigation.

**METHODS:** We performed RNA-seq analysis of RNA transcripts of human peripheral blood mononuclear cells from a case-control study including subjects with self-reported depression in the pre-symptomatic state of major depressive disorder and analyzed differentially expressed genes (DEGs) and the frequency of intron retention (IR) using rMATS.

**RESULTS:** Among the statistically significant DEGs identified, 651 upregulated and 820 downregulated genes were enriched in the Gene Ontology term ‘innate and adaptive immunity’. The upregulated DEGs were particularly enriched in the term ‘bacterial infection and phagocytosis’, whereas the downregulated DEGs were enriched in the terms ‘antigen presentation’ and ‘T-cell proliferation and maturation’. We also analyzed 158 genes for which IR was increased (IncIR) and 211 genes for which IR was decreased (DecIR) among the depressed subjects. The Gene Ontology terms associated with IncIR and DecIR were very similar to those of the up- and downregulated genes, respectively, with a preponderance of the term ‘ciliary assembly and function’ for DecIR. Moreover, the results of the network analysis also showed that a Japanese herbal medicine could partially mitigate the severity of depression among depressed patients. Inclusion of both IncIR and DecIR genes in the network analysis revealed several pathways related to the ability of patients to recover from depression.

**CONCLUSION:** Depression was found to be associated with activation of the innate immune response and relative inactivation of T-cell signaling. The DEGs we identified reflect physiological demands that are controlled at the transcriptional level, whereas the IR results reflect a more direct mechanism for monitoring protein homeostasis. Accordingly, an overall increase in IR is a stress response, and intron-retained transcripts are sensors of the physiological state of the cytoplasm. In particular, detection of increased IR in cilia-specific genes apparently correlates with defects in ciliary function or immunological synapse in depressed subjects. The results demonstrate the potential of relative IR as a biomarker for the immunological stratification of depressed patients and the utility of IR for the discovery of novel pathways involved in recovery from depression.

## Introduction

In 2017, it was estimated that more than 320 million people worldwide were affected by clinical depression (1). It is likely that this number—along with the number of people affected by other mental disorders—has since risen owing to ever-increasing stresses associated with daily life, particularly since the outbreak of COVID-19. In fact, depression has become the leading cause of disability worldwide. Major depressive disorder (MDD) is often accompanied by anxiety disorder, and this combination is the leading cause of death by suicide (1). Currently, there is no reliable laboratory test or effective treatment strategy to diagnose or cure MDD. Another important issue in depression is the low remission rate, with approximately one-half of patients achieving complete remission; moreover the remission rate decreases with each subsequent treatment. Therefore, to better understand the pathogenesis of depression and its etiology, there is urgent need to identify biomarkers for monitoring treatment outcomes as well as genes that can be targeted for drug therapy (2, 3, 4, 5, 6).

An increasing body of evidence suggests that the incidence of peripheral inflammation correlates with that of depression (7, 8, 9, 10, 11, 12, 13, 14). Several case-control studies of MDD patients have reported elevated peripheral-blood levels of inflammatory cytokines such as C-reactive protein, interleukin 6, and tumor necrosis factor (15, 16, 17). In these cases, for which inflammation occurs first and depressive symptoms appear later, the viewpoint that inflammation contributes to depression is gaining ground. Furthermore, the prevalence of depression as a comorbidity is rather high for many inflammatory diseases that do not have a mental-health comorbidity, such as rheumatoid arthritis (18), suggesting a possible role for inflammation in depression. In his excellent book “THE INFLAMED MIND” (19), Ed Bullmore proposed that stress causes inflammation and that inflammation causes depression. Much of the current data seem to be consistent with his proposal (20).

Alternative pre-mRNA splicing is a mechanism by which multiple protein isoforms can be produced from a single gene transcript. One type of alternative splicing is termed ‘intron retention’ (IR), which was previously thought to simply reflect one or more errors during pre-mRNA splicing. More recently, however, IR has been considered to be a biologically meaningful phenomenon, as an increase or decrease of intron abundance among specific transcripts has been associated with certain phenomena such as cell differentiation (21, 22, 23, 24), aging (25), and oncogenesis (26). Using mouse models of aging such as Klotho mice (27) and SAMP8 mice (28), we previously showed that the frequency of IR increases in response to stress in the pre-symptomatic state and that when the state is restored by administration of a Japanese herbal medicine, the incidence of IR is restored to that of the healthy state (27, 28). In addition, we proposed that genes affected by IR (herein termed IR genes) play a sensor role in detecting perturbations in cellular homeostasis (29).

We therefore hypothesized that an analysis of IR genes could facilitate the identification of stresses experienced by patients and the possibility that dysfunctional genes may underlie their depression. In this regard, we explored the possibility that the incidence of IR could be used to investigate the etiology of depression.

## Materials and Methods

### Ethics declarations, ethics approval and consent to participate

The research plan was reviewed and approved by the Research Ethics Committee of Kitasato Institute Hospital and assigned research number 21037. The study on which this research was based was an interventional study with the following approval numbers: No. 21039 and UMIN Study ID UMIN000045707. The Kitasato Institute Hospital Research Ethics Committee deliberates in accordance with the Ethics Guidelines for Medical and Health Research Involving Human Subjects in Japan. All participants gave written consent to the research procedures, including genetic analysis.

### Subjects

We recruited subjects with depressive symptoms who had consented to participate in the “Study of Hangekobokuto (30) and the Intestinal Environment” conducted by the Kitasato University Oriental Medicine Research Center and who scored between 6 and 20 on the Brief Depressive Symptom Scale. After the benefits and risks of the study were explained to each subject, written informed consent was obtained. For subjects who consented, we applied the following eight exclusion criteria. 1) Subjects already receiving drug treatment for depression; 2) subjects who had taken herbal medicinal preparations within the previous 4 weeks; 3) subjects who had taken antibiotics within the previous 4 weeks; 4) subjects who were clearly in need of conventional medical treatment; 5) subjects who had been diagnosed with ulcerative colitis or Crohn’s disease; 6) subjects with clinically significant hepatic or renal impairment; 7) subjects who had participated in other clinical studies within the past 12 weeks; and 8) subjects who were deemed by the investigators to be unsuitable for the study.

Each subject took a single daily dose of the Japanese herbal medicine Hangekobokuto (HKT) at home. The BDI™-II Beck Depression Questionnaire (BDI-II) was administered at the time of initial screening and at hospital visits 2 months after taking their final dose of HKT, and blood was collected using a BD Vacutainer CPTTM Blood Collection Tube (Nippon Becton Dickinson, Japan). Subjects were classified according to their BDI-II score, with six subjects scoring less than 16 and being considered as controls (CON) and eight subjects with depression symptoms (before medical treatment, BMT, or after medical treatment, AMT) scoring 17 or higher.

### Japanese herbal medicine

Japanese herbal medicines originated in ancient China and are widely used in Japan for treatment of a variety of conditions (31, 32). HKT (30) is one such formulation and is taken for symptoms of anxiety, stagnant gas in the stomach, and poor digestive function. In this study, HKT was used as a decoction in the following amounts, based on the formula of the Kitasato University Oriental Medicine Research Centre: Hange (Pinelliae Tuber) 6.0 g; Bukuryo (Hoelen) 5.0 g; Koboku (Magnoliae Cortex) 3.0 g, Shisoyo (Perillae Herba) 2.0 g; Syokyo (Zingiberis Rhizoma) 0.5 g.

### Preparation of PBMCs, RNA extraction, RNA library preparation, and RNA sequencing (RNA-seq)

Blood samples were centrifuged within 2 h to isolate the PBMC (peripheral blood mononuclear cell) layer. After centrifugation, PBMC samples were stored at –80 °C. RNA was extracted from individual PBMC samples. Library construction and paired-end sequencing (150 base pairs × 2) using the NovaSeq 6000 platform (Illumina) were outsourced to Azenta Life Sciences, Tokyo, Japan. RNA-seq yielded 109–148 million (× 2, paired-ends) raw reads per sample. These were then narrowed down using conventional procedures (33, 34, 35, 36).

### Analysis of differentially expressed genes

Using the edgeR package in R, significantly differentially expressed genes (DEGs) in patients with depression were detected by performing the likelihood ratio test. The results showed that 922 downregulated and 641 upregulated genes were significantly differentially expressed between the six CONs and eight BMTs (P < 0.05 and fold-change > 1.2). DEGs were used for GO (Gene Ontology) and KEGG (Kyoto Encyclopedia of Genes and Genomes) pathway-enrichment analysis using the DAVID website. Similarly, the same likelihood ratio test values were calculated under the same conditions between BMT and AMT to investigate the effect of HKT administration.

### Detection of IR

IR-containing genes were analyzed to determine their possible role in stress sensing as proposed previously (27, 28, 29). rMATS v.4.1. was used to assess the differential IR landscape embedded in the RNA-seq data. For our analysis, the parameters for the rMATS program were as follows: [--cstat 0.05 -t paired --readLength 150 --variable-read-length]. A cut-off of P < 0.05 in the likelihood ratio test and an absolute difference of the IR ratio > 0.05 (both used to establish statistical significance in the rMATS program) were used to call differential IR events. Similarly, the same likelihood ratio test values were calculated under the same conditions between BMT and AMT to study the effect of HKT administration.

### Interactome analysis

A protein-protein interaction network was generated using Cytoscape ver. 3.9.1 with StringApp version 1.7.1. The network type “full STRING network” was selected for plotting graphs, and a confidence-score cut-off value of 0.7 was used (default values were used for all other parameters). The statistical significance of data presented in Figure 5 was established using proteins encoded by IR genes and DEGs, and protein-protein interactions were analyzed among members of each functional gene group, cilia-related genes (proteins), psychiatric disorders–relevant genes (proteins), and adaptive and innate immunity–related genes (proteins).

## Results

### RNA-seq and analysis of DEGs

The study included a group of eight subjects with depression for whom the BDI-II score ranged from 17 to 27, which reflects moderate depression. These subjects were designated as the BMT group. Six other subjects with relatively mild depression had scores ranging from 7 to 16, and these subjects served as controls (CON group). (see Figure 1A(i)(ii)). As we are all on the spectrum, we wanted to gain molecular insight into the transition from mild to moderate depression. All subjects were also screened to ensure that they had not taken any medication or been hospitalized during the 3-month period before examination (see Materials and Methods for details).

**Fig. 1.**
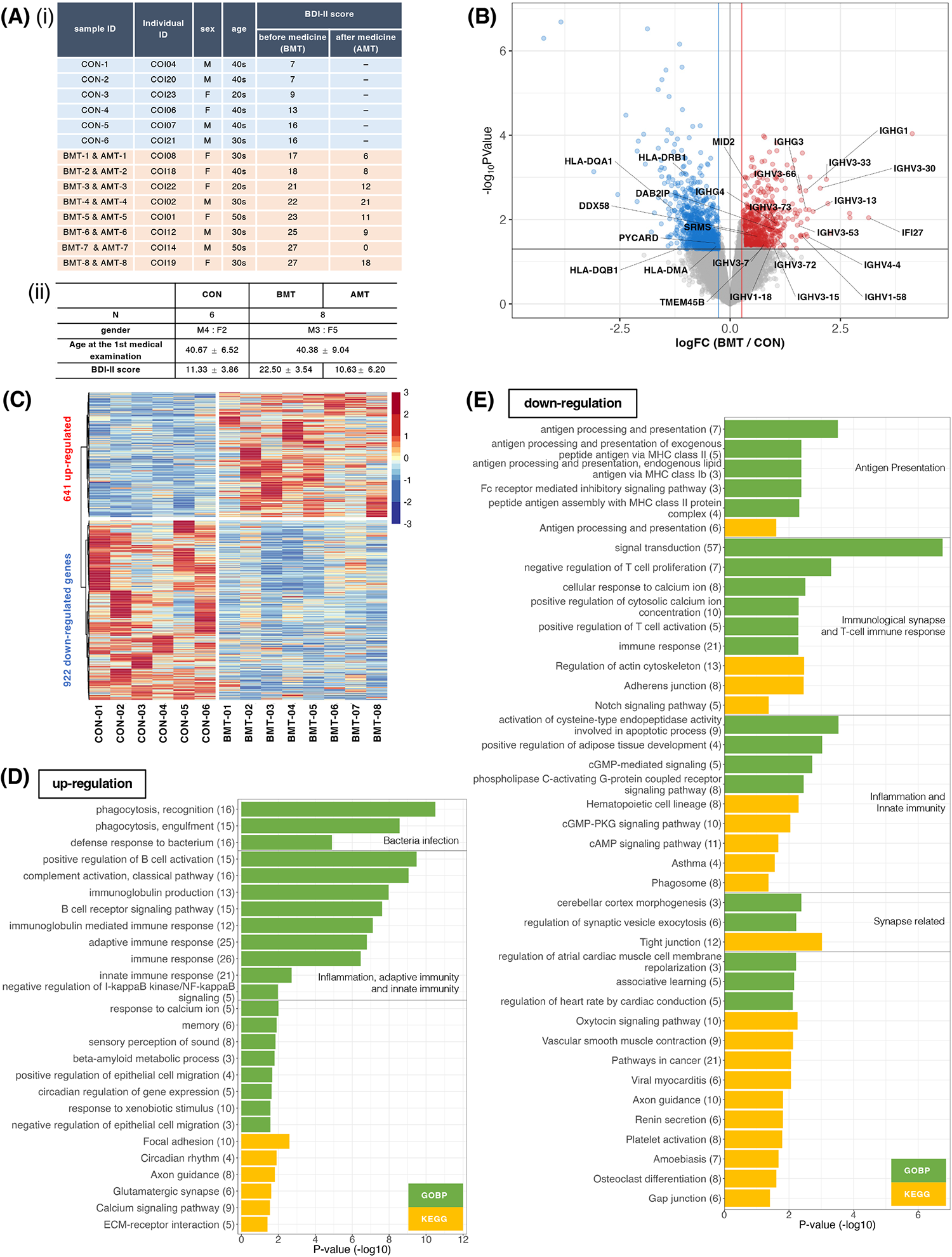
Comparison of RNA expression between depressed subjects and less depressed controls. (A) (ⅰ) Subject information. Classification was based on the BDI-II score at first examination. Subjects with BDI-II score < 16 were categorized as less depressed (controls, CON), and those with BDI-II score ≥ 16 were categorized as depressed. BMT, depressed subjects before medical treatment; AMT, depressed subjects after 2 months of taking HKT. (ii) Mean and standard deviation of sex, age, and BDI-II score for each group. (B) Volcano plot of RNA expression between the BMT and CON groups. The horizontal axis shows the log2 fold-change of BMT/CON, and the vertical axis shows –log10 P-values. Red dots denote significantly upregulated genes (FC (fold change) > 1.2 and P < 0.05), blue dots denote significantly downregulated genes (FC > 1/1.2 and P < 0.05), and grey dots indicate no significant difference in expression (likelihood ratio test). Gene symbols for T cell–associated genes are indicated. (C) Heatmap of significantly differentially expressed genes between BMT and CON subjects. (D, E) Enrichment analysis of biological processes among Gene Ontology and KEGG pathway terms for the 641 upregulated (D) or 922 downregulated (E) genes in the BMT group. The horizontal axis shows –log10 P-values. Green bars indicate Gene Ontology biological process terms, and yellow bars indicate KEGG pathway terms.

PBMC and RNA were isolated from each sample and used for RNA-seq. DEG analysis resulted in the identification of 651 upregulated and 820 downregulated genes for the BMT group compared with the CON group (Figure 1BC, Supplementary Table 1). GO enrichment analysis of the upregulated genes (Figure 1D, Supplementary Figure 1) revealed enrichment for innate immunity–related terms such as infection, phagocytosis, inflammation and adaptive immunity– related terms; the analysis of downregulated genes (Figure 1E, Supplementary Figure 2) revealed enrichment for adaptive immunity–related terms such as antigen presentation, T-cell activation, and synapse-related terms. Figure 1B shows all downregulated genes involved in T-cell activation (5 genes in Figure 1E) and all upregulated genes involved in the innate immune response (21 genes in Figure 1D), which included a large number of immunoglobulin heavy chains (37, 38), suggesting that many of the subjects had an inflammatory phenotype (37).

### Identification of IncIR and DecIR genes in depressed subjects

Because IR is a stress response and genes susceptible to IR are a physiological sensor ((27, 28); see later), we characterized genes for which IR was increased in depressed subjects (IncIR) and genes for which IR was decreased (DecIR), considering that such an analysis would indicate the type of stress to which the subjects had been exposed. A total of 158 IncIR and 198 DecIR genes were identified (Figure 2AB, Supplementary Table 2), and their characteristics were first studied based on published information. As expected, many sensor or regulatory genes were represented among the protein-coding IR genes (Table 1; 45 genes). This is only half of the genes identified as sensors, regulators and modulators among the IR genes in this analysis, in which genes controlling inflammation, innate immunity and adaptive immunity were also identified. GO enrichment analysis (Figure 2CD) of IncIR revealed enrichment of terms related to the TNF signaling pathway and several terms related to innate immune response. In IncIR, the TNF signaling pathway and several terms related to innate immune response were enriched; among the DecIR genes, there was enrichment of terms related to T-cell signaling and other adaptive immune responses as well as inflammation and innate immune processes. In short, both innate and adaptive immunity were highlighted in the IR analysis, as was the case for the DEG analysis, suggesting that the IR genes mirrored the DEG genes. The important difference between the IR genes and DEG genes is that immunoglobulin genes were included in the DEG list—in fact, almost half of the upregulated genes in our RNA-seq analysis were immunoglobulin genes (see Supplementary Figure 1), but this was not true for the IR [genes (see Discussion). The reason why immunoglobulin genes were not among the IR genes is discussed later. Interestingly, the highest enrichment score among the DecIR genes was for cilium assembly, suggesting that cilia are involved in sensing depression-induced stress (see Discussion).

**Fig. 2.**
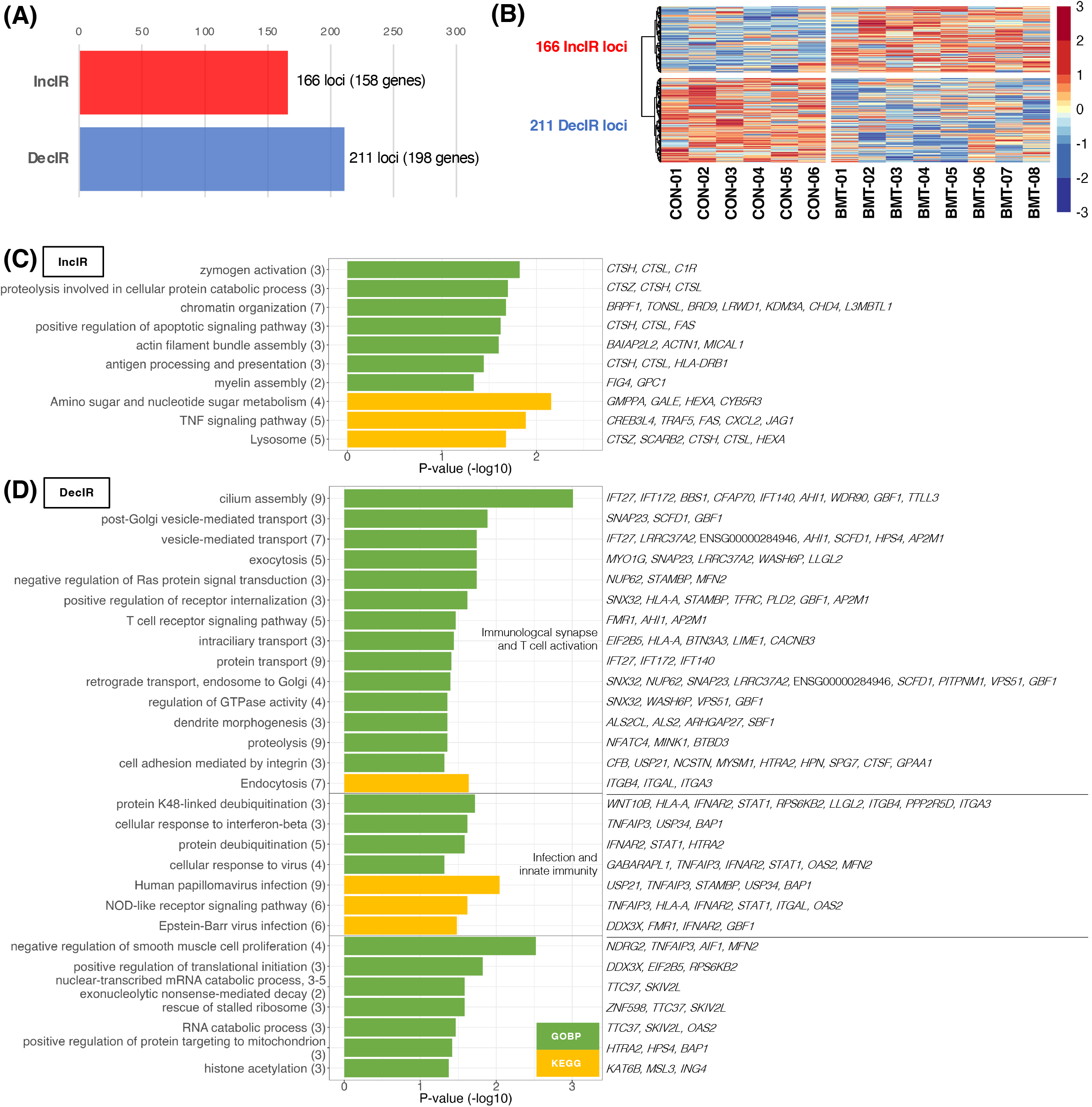
Identification and characterization of IR genes. (A) Number of IR genes with significantly increased IR (IncIR) or decreased IR (DecIR) for BMT versus CON using rMATS software v.4.1.1. The statistical significance of differences between values was based on a P-value of <0.05 and a difference in intron ratio of >0.05. (B) Heatmap of significantly different IR loci between BMT and CON. (C, D) Enrichment analysis of biological processes among Gene Ontology and KEGG pathway terms using 158 IncIR genes (C) and 198 DecIR genes (D) in the BMT group. Gene symbols corresponding to the terms are shown on the right. The horizontal axis shows –log10 P-values. Green bars indicate Gene Ontology biological process terms, and yellow bars indicate KEGG pathway terms.

**Table 1.**
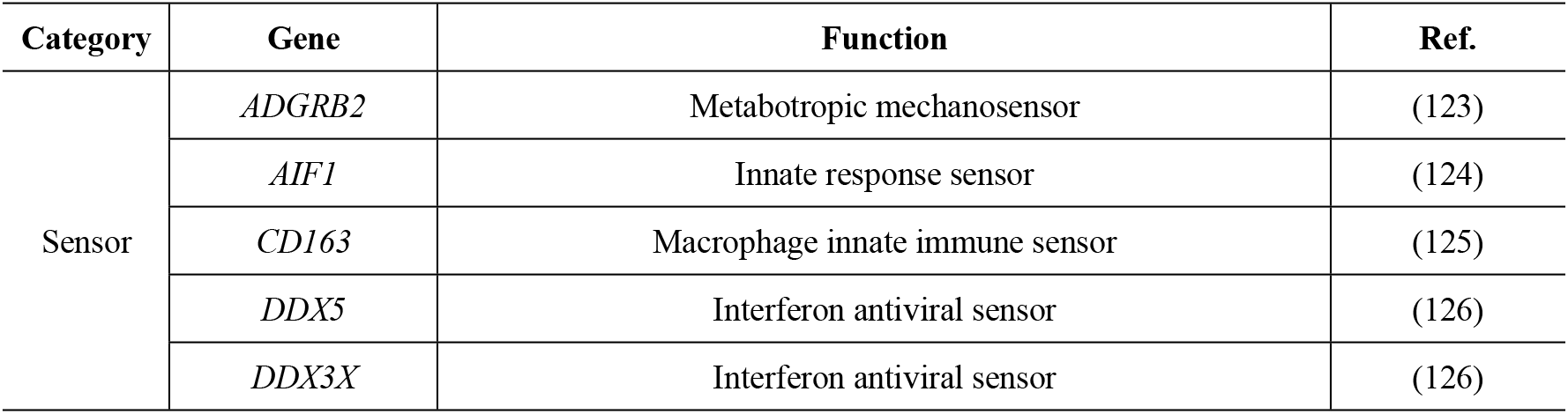
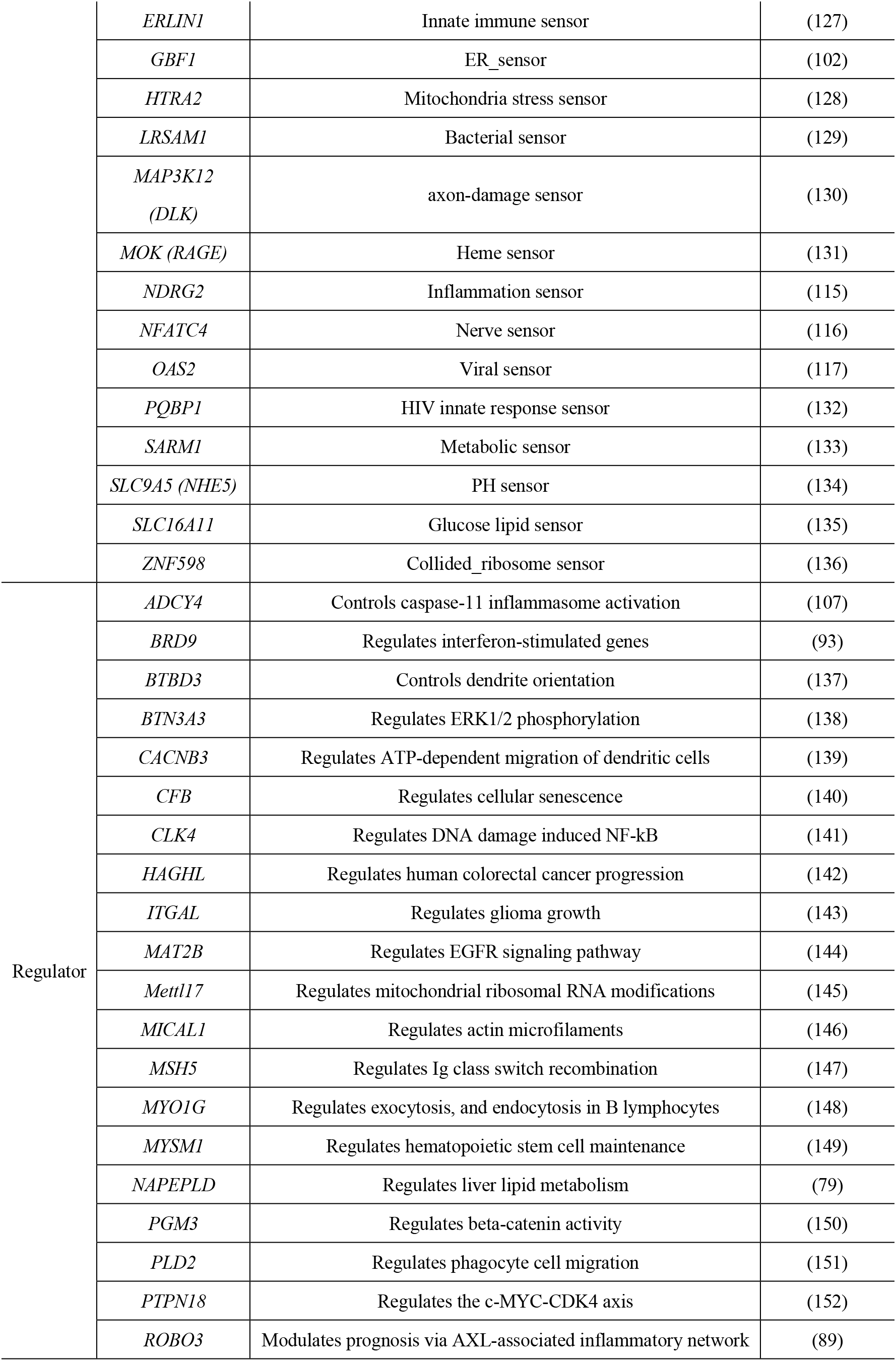
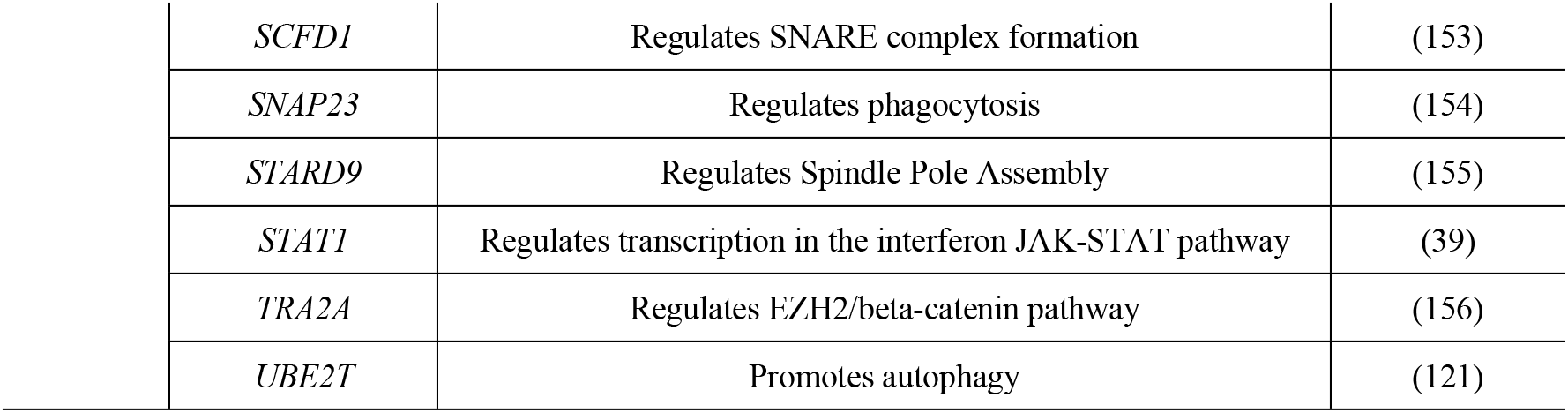
Sensor and regulatory genes that were identified from among the protein-coding IR genes.

| Category | Gene | Function | Ref. |
| --- | --- | --- | --- |
| Sensor | *ADGRB2* | Metabotropic mechanosensor | (123) |
|  | *AIF1* | Innate response sensor | (124) |
|  | *CD163* | Macrophage innate immune sensor | (125) |
|  | *DDX5* | Interferon antiviral sensor | (126) |
|  | *DDX3X* | Interferon antiviral sensor | (126) |
|  | *ERLIN1* | Innate immune sensor | (127) |
|  | *GBF1* | ER_sensor | (102) |
|  | *HTRA2* | Mitochondria stress sensor | (128) |
|  | *LRSAM1* | Bacterial sensor | (129) |
|  | *MAP3K12*<br>(*DLK*) | axon-damage sensor | (130) |
|  | *MOK (RAGE)* | Heme sensor | (131) |
|  | *NDRG2* | Inflammation sensor | (115) |
|  | *NFATC4* | Nerve sensor | (116) |
|  | *OAS2* | Viral sensor | (117) |
|  | *PQBP1* | HIV innate response sensor | (132) |
|  | *SARM1* | Metabolic sensor | (133) |
|  | *SLC9A5 (NHE5)* | PH sensor | (134) |
|  | *SLC16A11* | Glucose lipid sensor | (135) |
|  | *ZNF598* | Collided_ribosome sensor | (136) |
| Regulator | *ADCY4* | Controls caspase-11 inflammasome activation | (107) |
|  | *BRD9* | Regulates interferon-stimulated genes | (93) |
|  | *BTBD3* | Controls dendrite orientation | (137) |
|  | *BTN3A3* | Regulates ERK1/2 phosphorylation | (138) |
|  | *CACNB3* | Regulates ATP-dependent migration of dendritic cells | (139) |
|  | *CFB* | Regulates cellular senescence | (140) |
|  | *CLK4* | Regulates DNA damage induced NF-kB | (141) |
|  | *HAGHL* | Regulates human colorectal cancer progression | (142) |
|  | *ITGAL* | Regulates glioma growth | (143) |
|  | *MAT2B* | Regulates EGFR signaling pathway | (144) |
|  | *Mettl17* | Regulates mitochondrial ribosomal RNA modifications | (145) |
|  | *MICAL1* | Regulates actin microfilaments | (146) |
|  | *MSH5* | Regulates Ig class switch recombination | (147) |
|  | *MYO1G* | Regulates exocytosis, and endocytosis in B lymphocytes | (148) |
|  | *MYSM1* | Regulates hematopoietic stem cell maintenance | (149) |
|  | *NAPEPLD* | Regulates liver lipid metabolism | (79) |
|  | *PGM3* | Regulates beta-catenin activity | (150) |
|  | *PLD2* | Regulates phagocyte cell migration | (151) |
|  | *PTPN18* | Regulates the c-MYC-CDK4 axis | (152) |
|  | *ROBO3* | Modulates prognosis via AXL-associated inflammatory network | (89) |

|  |  |  |
| --- | --- | --- |
| *SCFD1* | Regulates SNARE complex formation | (153) |
| *SNAP23* | Regulates phagocytosis | (154) |
| *STARD9* | Regulates Spindle Pole Assembly | (155) |
| *STAT1* | Regulates transcription in the interferon JAK-STAT pathway | (39) |
| *TRA2A* | Regulates EZH2/beta-catenin pathway | (156) |
| *UBE2T* | Promotes autophagy | (121) |

### IR genes interact with genes related to the innate immune response in a statistically significant manner

To characterize the IR genes in more detail, we first determined the possible overlap of IR genes with immune-related genes, cilia genes, and psychiatric disease–related genes (designated PD; Figure 3AB). The 317 IR genes (Supplementary Table 2) included 32 cilia genes (Figure 3C) and 34 immune-related genes (Figure 3D). Because many IR genes listed in Table 1 are involved in innate immunity, including viral and bacterial infection, we next examined whether IR genes could specifically interact with genes involved in the innate immune response (Figure 3E). Indeed, IR genes were found to interact specifically with genes involved in innate immunity (statistically significant, P < 0.0234). Figure 3F shows the ranking of the IR gene interactions. Among the IR genes, *STAT1* (signal transducer and activation of transcription gene 1; Table 1 (39)) had the greatest number of interactions with innate immunity genes. Notably, *STAT1* is involved in the JAK-STAT pathway (39), which contributes to both innate and adaptive immunity including inflammation (see Discussion). We also assessed interactions between IR genes and genes involved in leukocyte activation (adaptive immunity) or the immune response, revealing that *STAT1* ranked highest in each comparison (Figure 3JK).

**Fig. 3.**
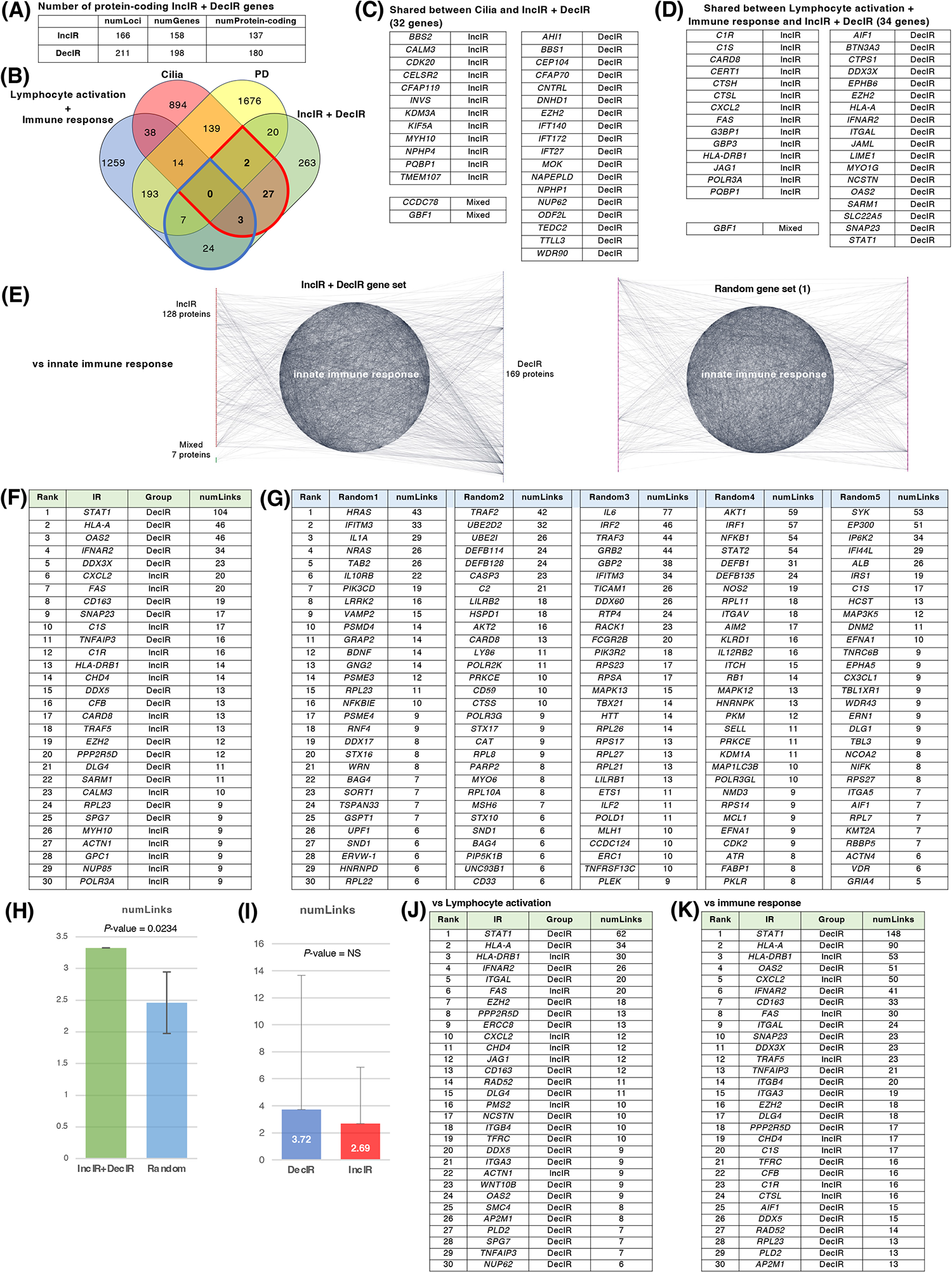
IR loci interact preferentially with genes involved in innate immunity. (A) Number of IR genes and IR protein-coding genes. (B) Venn diagram between IR genes, cilia-related genes (GO: 0060271 cilium assembly + SCGSv2 ciliary genes (69)), adaptive and innate immunity– related genes (GO: 0046649 lymphocyte activation and GO: 0006955 immune response), and psychiatric disorders–relevant (PD) genes. The PD gene set was constructed by merging genes from the following databases or previous studies: SFARI (autism-related gene database, https://www.sfari.org/resource/sfari-gene/); PsyGeNET (mental disorder-related gene database, https://www.sfari.org/resource/sfari-gene/); PD genes http://www.psygenet.org/web/PsyGeNET/menu/home); PD genes (70); major depression risk genes (71). (C) Gene symbols shared between IR and cilia-related genes, corresponding to the area outlined in red in B. Groups were classified as IncIR, DecIR or Mixed (’Mixed’ indicates a gene for which one intron was classified as IncIR and another classified as DecIR). (D) Gene symbols shared between IR and immunity–related genes, corresponding to the area outlined by blue in B. (E) Network showing significant protein-protein interactions (PPI) between proteins encoded by IR genes and those encoded by innate immune response–related genes. The interaction score was calculated using the full STRING network confidence score 0.7 from the STRING database. (Left) Network of PPIs between proteins encoded by IR genes and innate immune response genes (GO: 0045087). Innate immune-response proteins were placed in the central circle and IR proteins (red: IncIR, blue: DecIR, green: Mixed) on either side. (Right) Instead of IR proteins, equal numbers of randomly selected gene sets were placed on both sides. Ranking table shows the top 30 proteins with the greatest number of interactions (number of links) with the IR genes in (F) and with 5 randomly selected protein sets in (G). (H) Comparison of the average number of interactions (number of links) with innate immune-response proteins among the IR genes and 5 random protein sets. (I) Comparison of the average number of interactions (number of links) with innate immune-response proteins of genes classified as IncIR or DecIR. (J) Ranking table showing the top 30 proteins with the greatest number of interactions (links) to lymphocyte-activation proteins (GO: 0046649) among the IR proteins. (K) Ranking table showing the top 30 proteins with the greatest number of interactions (links) to immune-response proteins (GO: 0006955) among the IR proteins.

### IR-DEG interactome

Using all the protein-coding genes of the DEGs (285 upregulated + 433 downregulated) and IR genes (129 IncIR + 172 DecIR + 8 common to both), we created a large interactome (Figure 4A). Within the large interactome, many hub genes were connected to other genes (Supplementary Table 3). The largest hub centered on *SRC* (40), one of the DEGs, which was connected to 43 genes (Figure 4B). Among the IR genes, the largest hub was *DLG4* (41), which is involved in synaptic function. The second was *STAT1*. The third and fifth were integrin genes (42, 43), and the fourth was HLA-A (44), which is involved in antigen presentation. The sixth was MYH10 (45), myosin heavy chain, having 11 links, one of which is linked to myosin light chain kinase (MYLK) (46, 47); importantly, IR of the *MYH10* transcript was restored in subjects after administration of HKT (see Discussion).

**Fig. 4.**
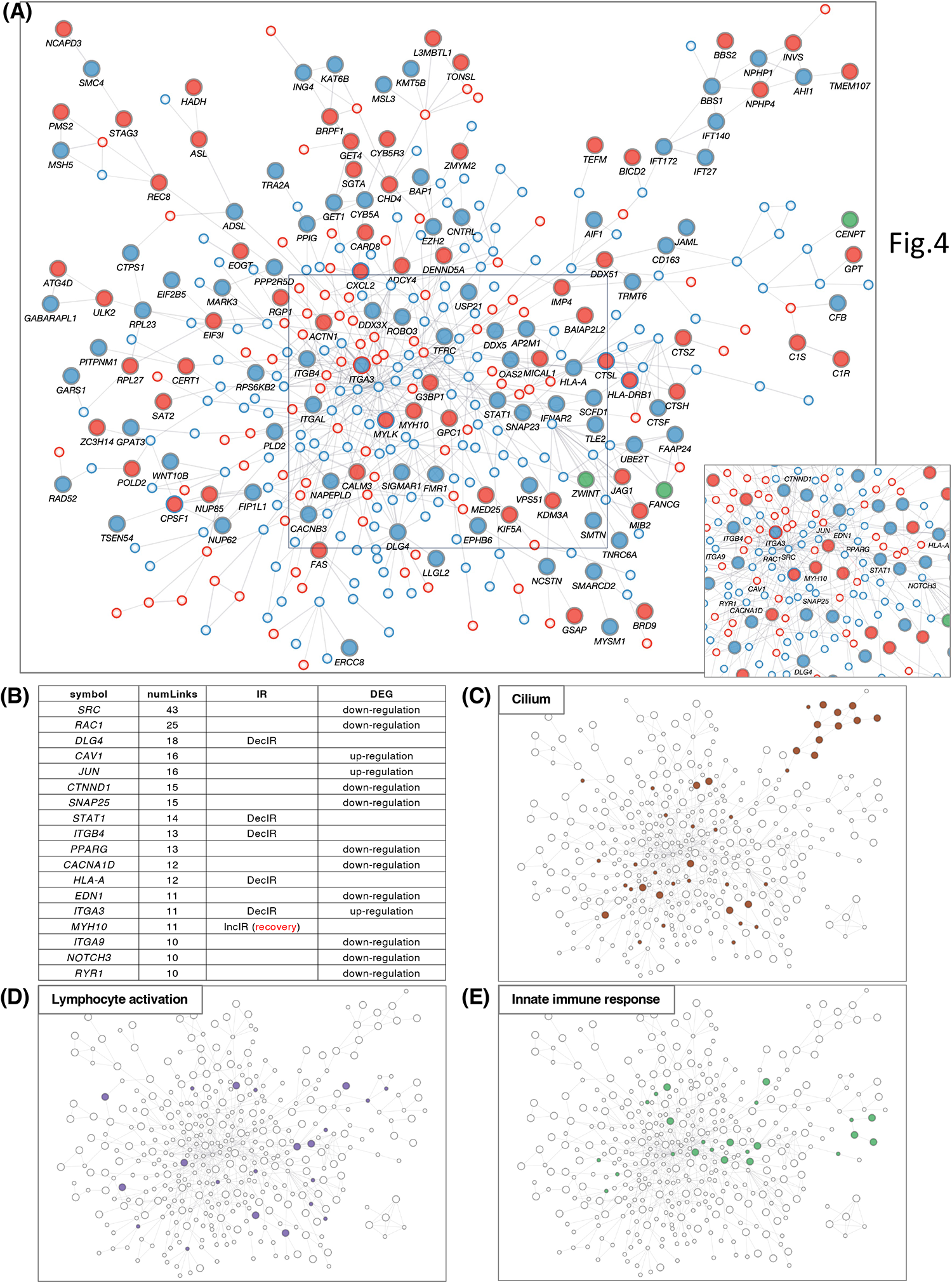
PPI network of IR and DEG proteins. (A) Main network showing PPIs using all IR and DEG proteins between BMT and CON. The interaction score was calculated using the full STRING network confidence score 0.7 from the STRING database. IncIR: red circles, DecIR: blue circles, Mixed: green circles. Upregulation: small circle with red border, downregulation: small circle with blue border. Only the largest networks are shown, as singletons and smaller networks were excluded. IR protein names are shown. Inset, bottom-right: (A square bottom right) Proteins with ≥10 interactions (number of links) are marked with symbols. (B) Ranking table of protein names with 10 or more interactions. (C–E) Proteins corresponding to cilia (brown, GO: 0060271 cilium assembly) in (C), lymphocyte activation (purple, GO: 0046649 lymphocyte activation) in (D), and innate immunity (light green, GO: 0045087 innate immune response) in (E) are colored as in the network in panel A.

### Recovery of IR genes by administration of HKT to subjects

After 2 months of HKT administration, PBMCs were isolated from blood samples of the subjects, and RNA-seq was performed (Figure 1A). We characterized two types of IR genes for which IR was recovered after HKT administration, namely reverse V-shape recovery in Figure 5A, comprising 30 protein-coding genes and V-shape recovery in Figure 5B, comprising 34 protein-coding genes (Supplementary Table 4). Because only 17 genes (7 for the V-shape recovery and 10 for the reverse V-shape recovery) were restored among those identified as DEGs (Figure 5D), the fact that IR was recovered after HKT treatment in four times as many IR genes as DEGs suggested that IR is superior to DEGs as a marker for evaluating the efficacy of a drug in the present case.

**Fig. 5.**
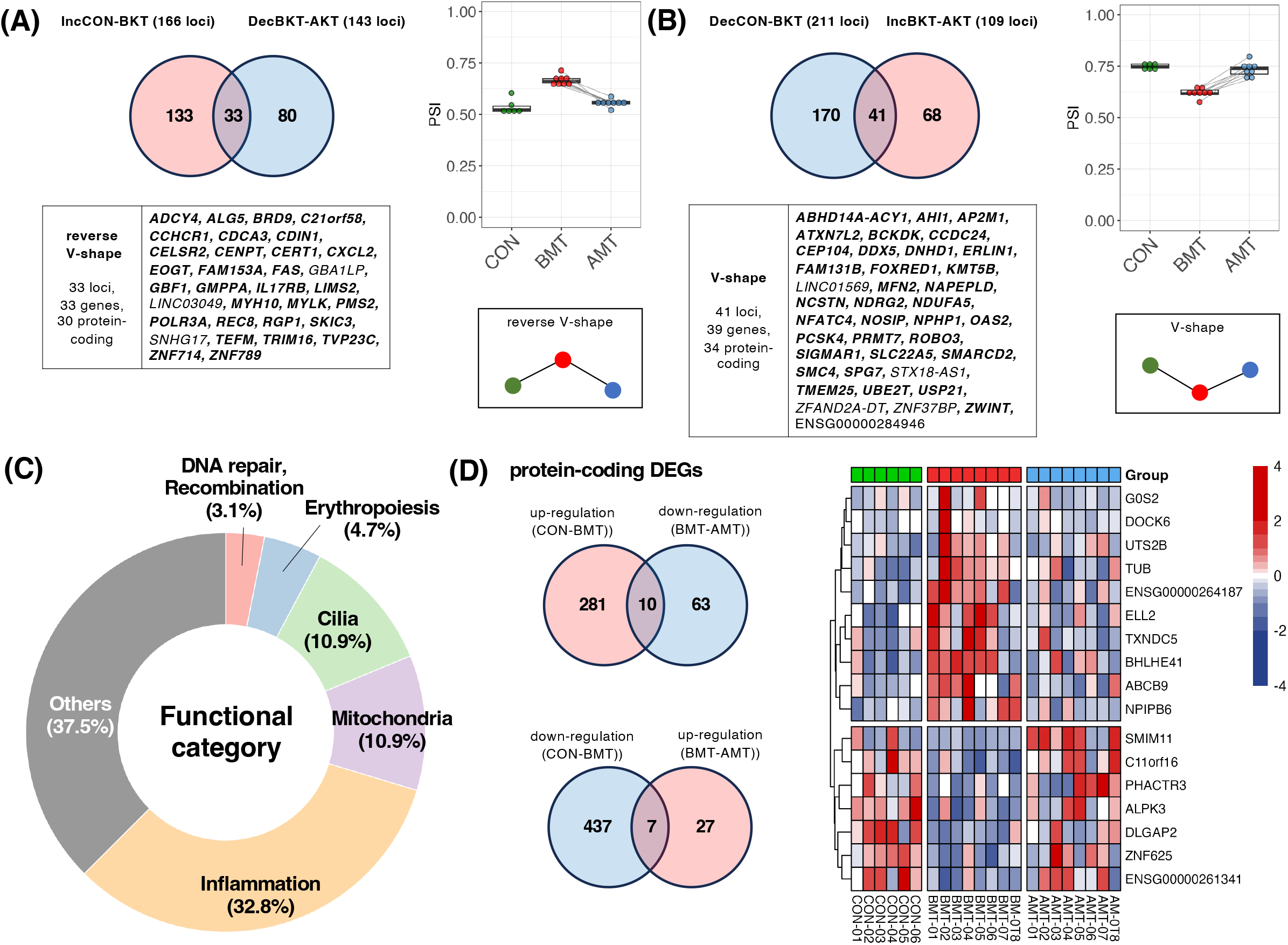
Recovery of IR by administration of HKT. (A) (Top, left) Venn diagram of intronic loci that were significantly increased (IncIR between BMT and CON) and those that were decreased (DecIR between BMT and AMT) by HKT treatment. (Bottom, left) Gene symbols with recovered loci are shown (protein-coding genes are shown in bold). (Right) Box plot showing average intron ratio at recovered loci. (B) (Top, left) Venn diagram of intronic loci that were significantly decreased (DecIR between BMT and CON) and those that were increased (IncIR between BMT and AMT) by HKT treatment. (Bottom, right) Gene symbols with recovered loci are shown (protein-coding genes are shown in bold). (C) Table of functional categorization from the literature of the recovered IR genes.

Characterization of the 64 protein-coding genes for which IR was restored after HKT treatment (Figure 5C) revealed that inflammation-related genes were the most affected (21 genes; 37.5%), with 7 mitochondria-related and 7 cilia-related genes—each accounting for 10.9%. The anti-inflammatory effect shown here is consistent with the reported efficacy of many Japanese herbal medicines (31), including HKT (30). Interestingly, whereas HKT has been reported to have anti-inflammatory effects by restoring the activity of inducible nitric oxide synthase, IR of the mRNA of the encoding gene, *NOSIP* (48), which modulates cellular NO level, was consistently recovered by HKT treatment (Figure 5B, Table 2). The identification of three hematopoiesis genes may also indicate that inflammation and hematopoiesis are linked in a compensatory way, as inflammation consumes a large number of macrophages, which should be supplied again. Oxidative stress is common in depressed patients (49) and can lead to increased DNA damage together with mitochondrial dysfunction (49). The restoration of these genes may be one of the hallmarks of this herbal medicine.

**Table 2.**
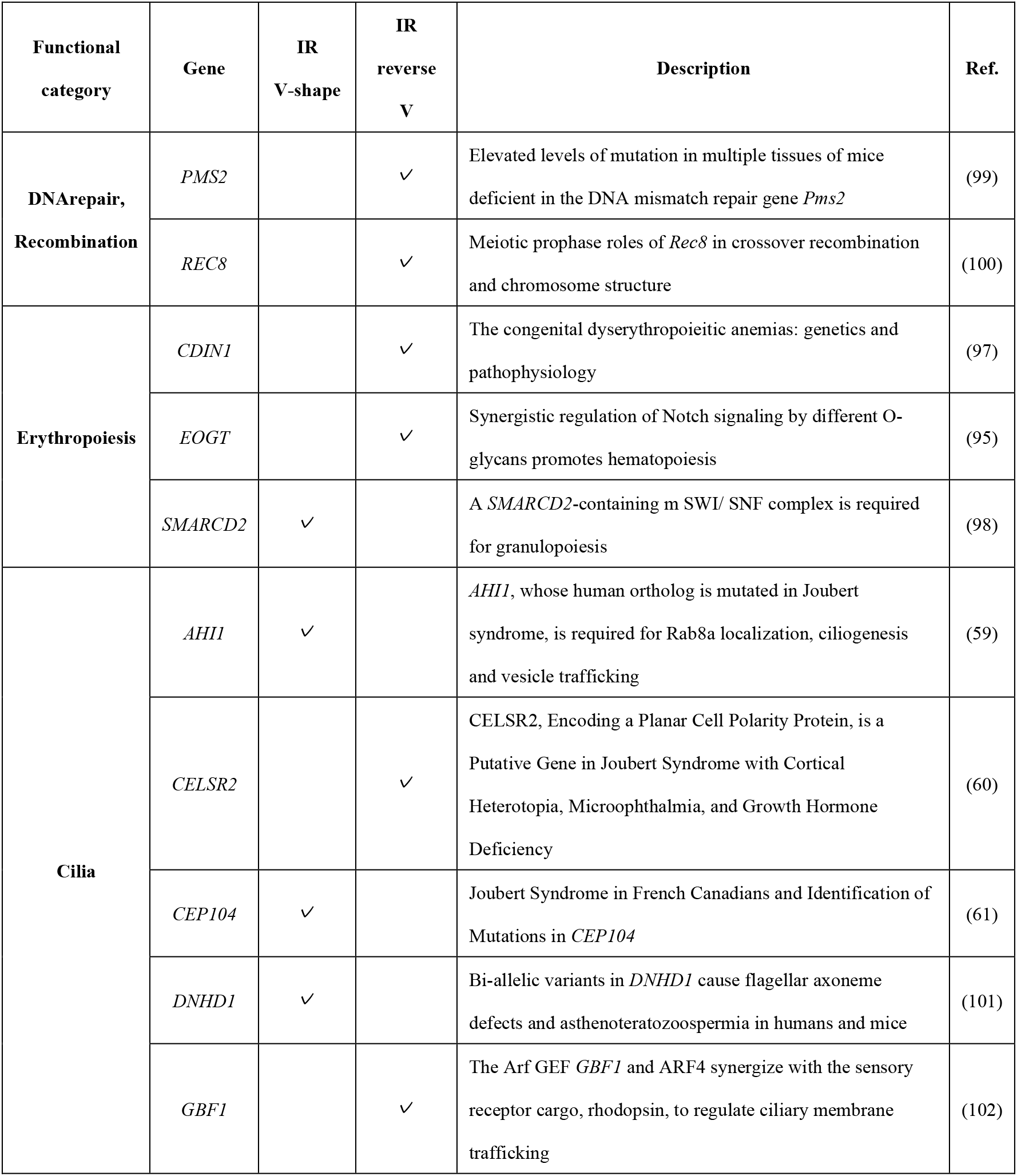
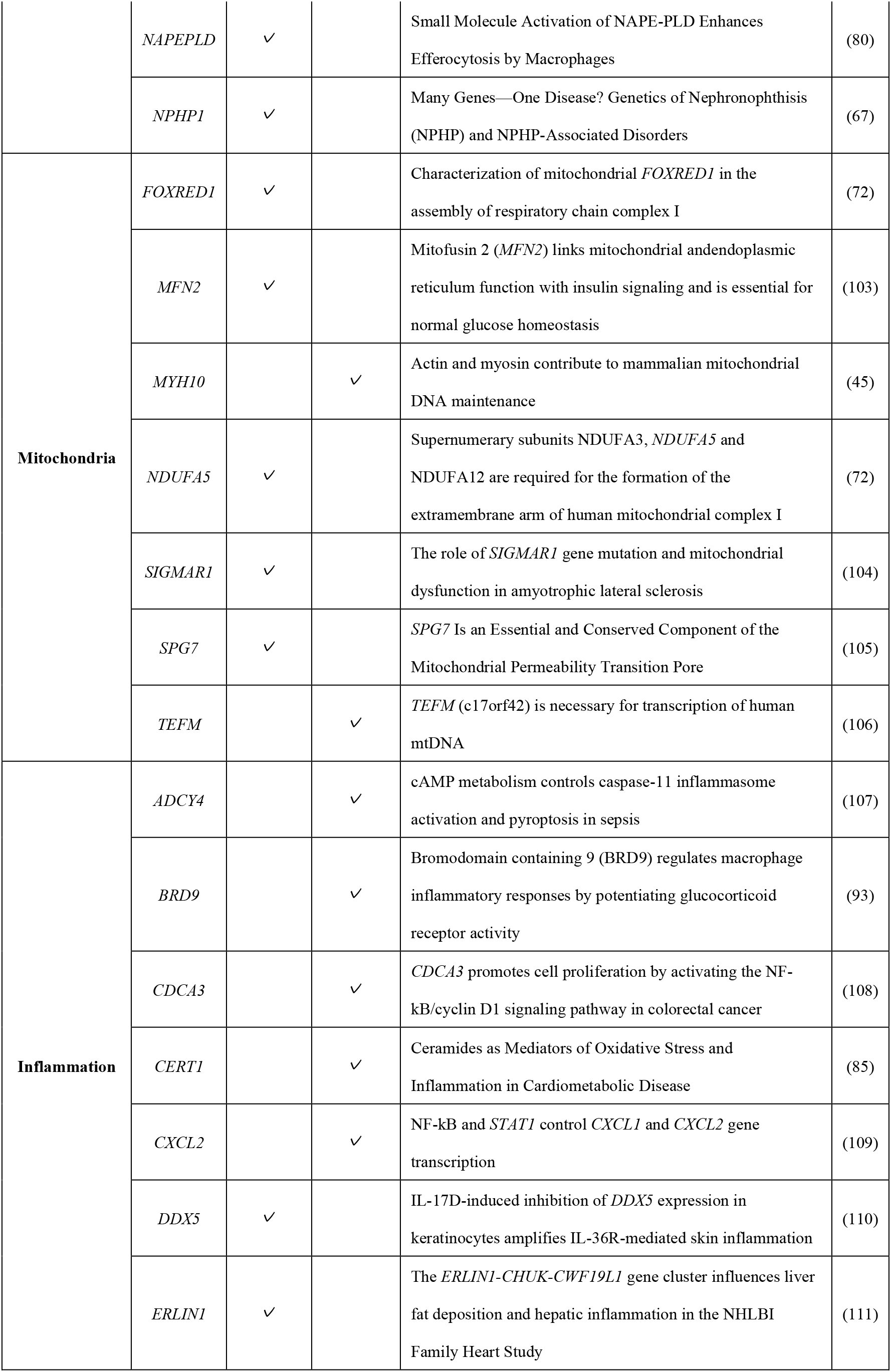
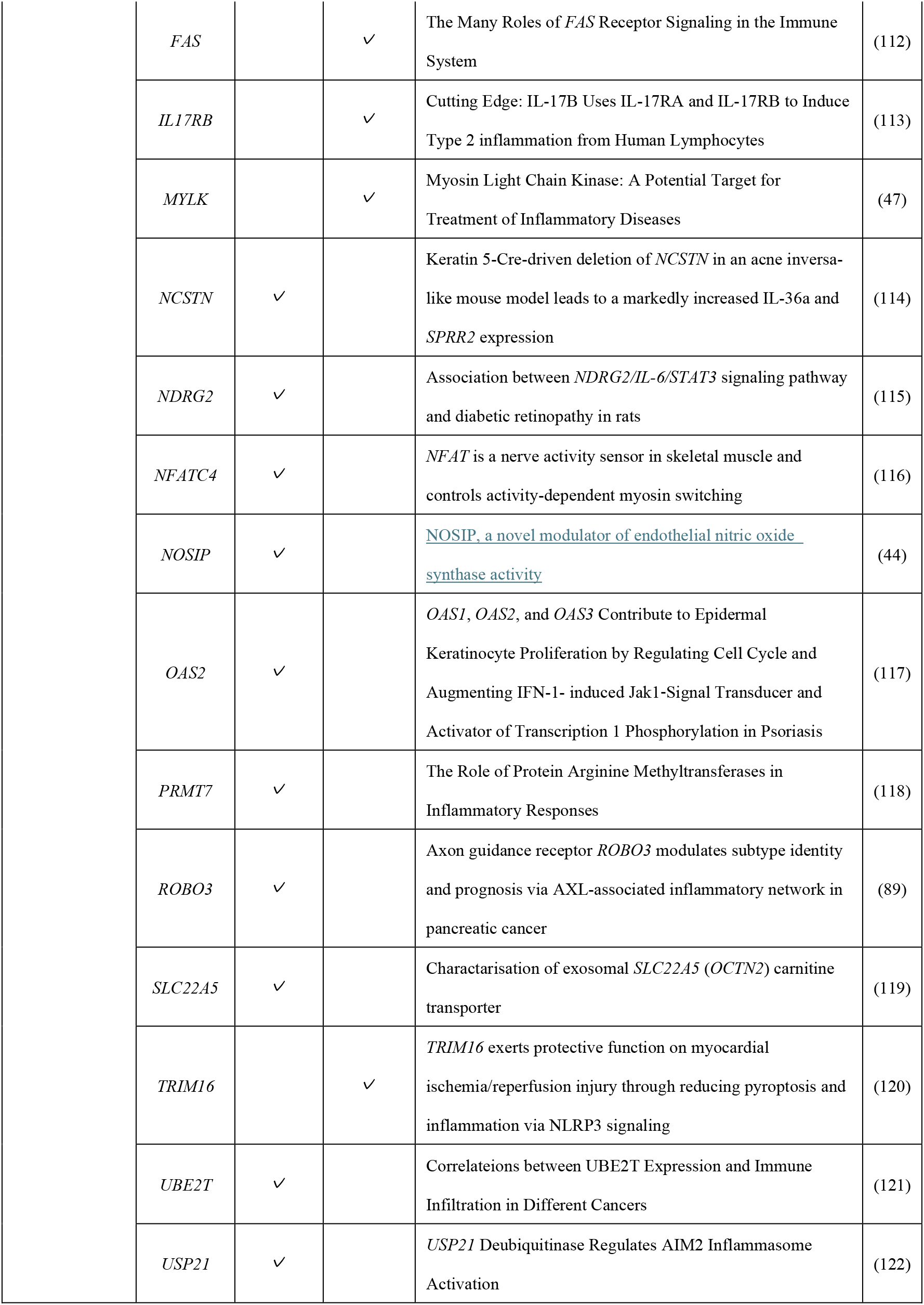
Functional categorization of recovery IR genes.

| Functional category | Gene | IR V-shape | IR reverse V | Description | Ref. |
| --- | --- | --- | --- | --- | --- |
| **DNA repair, Recombination** | *PMS2* |  | ✓ | Elevated levels of mutation in multiple tissues of mice deficient in the DNA mismatch repair gene *Pms2* | (99) |
|  | *REC8* |  | ✓ | Meiotic prophase roles of *Rec8* in crossover recombination and chromosome structure | (100) |
| **Erythropoiesis** | *CDIN1* |  | ✓ | The congenital dyserythropoietic anemias: genetics and pathophysiology | (97) |
|  | *EOGT* |  | ✓ | Synergistic regulation of Notch signaling by different O-glycans promotes hematopoiesis | (95) |
|  | *SMARCD2* | ✓ |  | A *SMARCD2*-containing m SWI/ SNF complex is required for granulopoiesis | (98) |
| **Cilia** | *AHI1* | ✓ |  | *AHI1*, whose human ortholog is mutated in Joubert syndrome, is required for Rab8a localization, ciliogenesis and vesicle trafficking | (59) |
|  | *CELSR2* |  | ✓ | CELSR2, Encoding a Planar Cell Polarity Protein, is a Putative Gene in Joubert Syndrome with Cortical Heterotopia, Microphthalmia, and Growth Hormone Deficiency | (60) |
|  | *CEP104* | ✓ |  | Joubert Syndrome in French Canadians and Identification of Mutations in *CEP104* | (61) |
|  | *DNHD1* | ✓ |  | Bi-allelic variants in *DNHD1* cause flagellar axoneme defects and asthenoteratozoospermia in humans and mice | (101) |
|  | *GBF1* |  | ✓ | The Arf GEF *GBF1* and ARF4 synergize with the sensory receptor cargo, rhodopsin, to regulate ciliary membrane trafficking | (102) |
|  | *NAPEPLD* | ✓ |  | Small Molecule Activation of NAPE-PLD Enhances Efferocytosis by Macrophages | (80) |
|  | *NPHP1* | ✓ |  | Many Genes—One Disease? Genetics of Nephronophthisis (NPHP) and NPHP-Associated Disorders | (67) |
| **Mitochondria** | *FOXRED1* | ✓ |  | Characterization of mitochondrial *FOXRED1* in the assembly of respiratory chain complex I | (72) |
|  | *MFN2* | ✓ |  | Mitofusin 2 (*MFN2*) links mitochondrial and endoplasmic reticulum function with insulin signaling and is essential for normal glucose homeostasis | (103) |
|  | *MYH10* |  | ✓ | Actin and myosin contribute to mammalian mitochondrial DNA maintenance | (45) |
|  | *NDUFA5* | ✓ |  | Supernumerary subunits *NDUFA3*, *NDUFA5* and *NDUFA12* are required for the formation of the extramembrane arm of human mitochondrial complex I | (72) |
|  | *SIGMAR1* | ✓ |  | The role of *SIGMAR1* gene mutation and mitochondrial dysfunction in amyotrophic lateral sclerosis | (104) |
|  | *SPG7* | ✓ |  | *SPG7* Is an Essential and Conserved Component of the Mitochondrial Permeability Transition Pore | (105) |
|  | *TEFM* |  | ✓ | *TEFM* (c17orf42) is necessary for transcription of human mtDNA | (106) |
| **Inflammation** | *ADCY4* |  | ✓ | cAMP metabolism controls caspase-11 inflammasome activation and pyroptosis in sepsis | (107) |
|  | *BRD9* |  | ✓ | Bromodomain containing 9 (BRD9) regulates macrophage inflammatory responses by potentiating glucocorticoid receptor activity | (93) |
| | *CDCA3* | | ✓ | *CDCA3* promotes cell proliferation by activating the NF-κB/cyclin D1 signaling pathway in colorectal cancer | (108) |
|  | *CERT1* |  | ✓ | Ceramides as Mediators of Oxidative Stress and Inflammation in Cardiometabolic Disease | (85) |
| | *CXCL2* | | ✓ | NF-κB and *STAT1* control *CXCL1* and *CXCL2* gene transcription | (109) |
|  | *DDX5* | ✓ |  | IL-17D-induced inhibition of *DDX5* expression in keratinocytes amplifies IL-36R-mediated skin inflammation | (110) |
|  | *ERLIN1* | ✓ |  | The *ERLIN1-CHUK-CWF19L1* gene cluster influences liver fat deposition and hepatic inflammation in the NHLBI Family Heart Study | (111) |

|  |  |  |  |  |
| --- | --- | --- | --- | --- |
| *FAS* |  | ✓ | The Many Roles of *FAS* Receptor Signaling in the Immune System | (112) |
| *IL17RB* |  | ✓ | Cutting Edge: IL-17B Uses IL-17RA and IL-17RB to Induce Type 2 inflammation from Human Lymphocytes | (113) |
| *MYLK* |  | ✓ | Myosin Light Chain Kinase: A Potential Target for Treatment of Inflammatory Diseases | (47) |
| *NCSTN* | ✓ |  | Keratin 5-Cre-driven deletion of *NCSTN* in an acne inversa-like mouse model leads to a markedly increased IL-36a and *SPRR2* expression | (114) |
| *NDRG2* | ✓ |  | Association between *NDRG2/IL-6/STAT3* signaling pathway and diabetic retinopathy in rats | (115) |
| *NFATC4* | ✓ |  | *NFAT* is a nerve activity sensor in skeletal muscle and controls activity-dependent myosin switching | (116) |
| *NOSIP* | ✓ |  | NOSIP, a novel modulator of endothelial nitric oxide synthase activity | (44) |
| *OAS2* | ✓ |  | *OAS1*, *OAS2*, and *OAS3* Contribute to Epidermal Keratinocyte Proliferation by Regulating Cell Cycle and Augmenting IFN-1- induced Jak1-Signal Transducer and Activator of Transcription 1 Phosphorylation in Psoriasis | (117) |
| *PRMT7* | ✓ |  | The Role of Protein Arginine Methyltransferases in Inflammatory Responses | (118) |
| *ROBO3* | ✓ |  | Axon guidance receptor *ROBO3* modulates subtype identity and prognosis via AXL-associated inflammatory network in pancreatic cancer | (89) |
| *SLC22A5* | ✓ |  | Characterisation of exosomal *SLC22A5* (*OCTN2*) carnitine transporter | (119) |
| *TRIM16* |  | ✓ | *TRIM16* exerts protective function on myocardial ischemia/reperfusion injury through reducing pyroptosis and inflammation via NLRP3 signaling | (120) |
| *UBE2T* | ✓ |  | Correlateions between UBE2T Expression and Immune Infiltration in Different Cancers | (121) |
| *USP21* | ✓ |  | *USP21* Deubiquitinase Regulates AIM2 Inflammasome Activation | (122) |

### Pathways for which IR was restored by HKT can be characterized from the DEG-IR interactome

The 64 protein-coding IR genes (Figure 5AB) and 17 DEGs (Figure 5D) for which IR was restored by HKT were overlaid on the DEG-IR interactome (shown in Figure 4) to determine whether any of these 81 genes could interact and network with each other. Ten new pathways were found (Figure 6). Protein-protein interactions in some of these pathways are known, but to our knowledge this is the first time that these pathways have been shown to be involved in restoring a physiological state. The implications of some of these pathways for the efficacy of HKT are discussed in the later section. As the IR-DEG interactome was generated independently of HKT treatment, this method offers the possibility that new pathways will be discovered when different drugs are used in similar subjects. Thus, IR-DEG interactome analysis could allow us to uncover new pathways involved in the mechanism of action of different types of drugs, including herbal medicines, and identify their commonalities and unique characteristics.

**Fig. 6.**
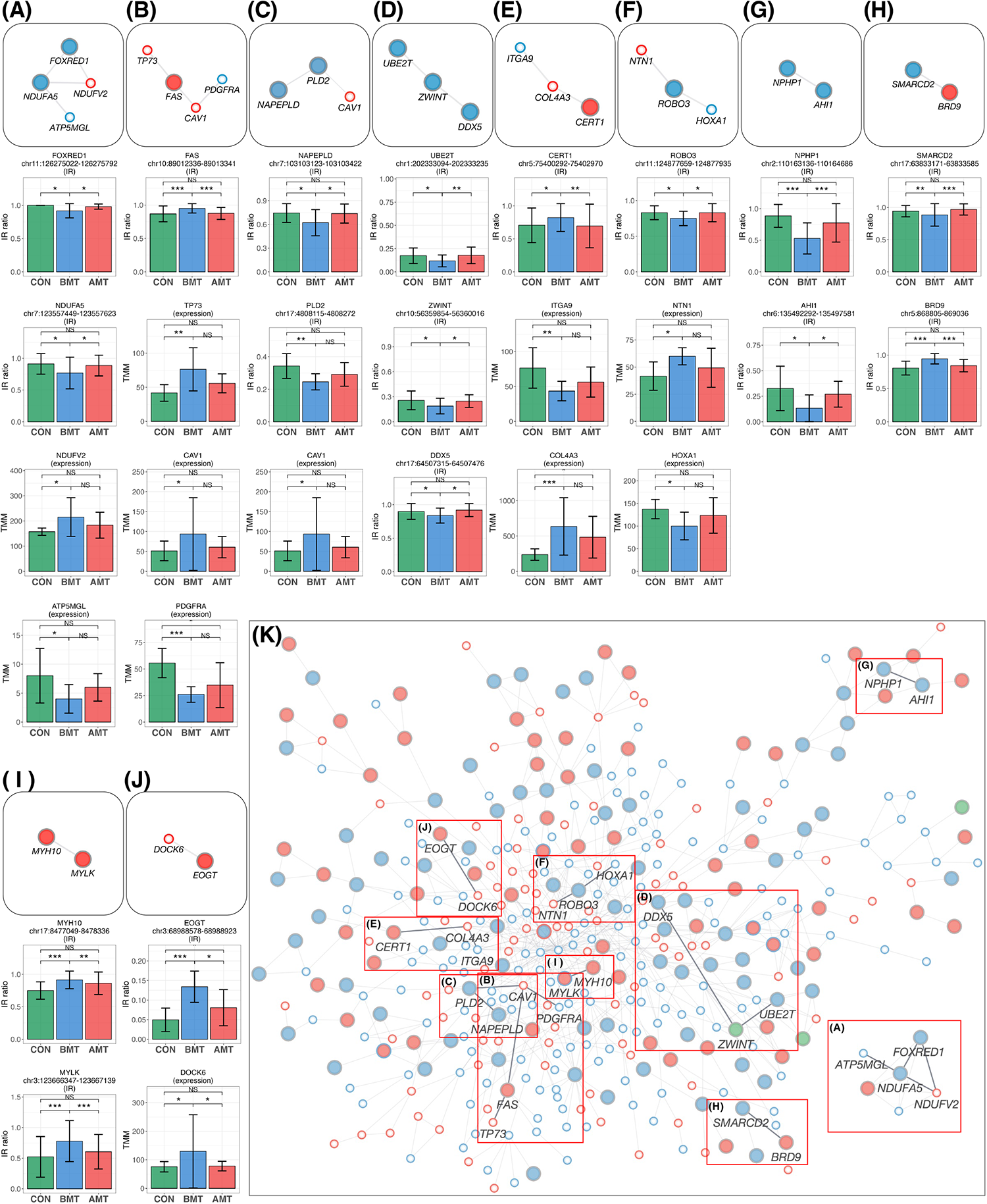
New pathways restored by HKT overlaid on the PPI network. (A–J) The network was extracted from the PPI network and then overlaid with the recovered IR and DEG loci. All recovered IR loci except *PLD2* showed a significant difference (P < 0.05, FC > 1.2) between BMT and CON and between BMT and AMT. All DEG loci showed a significant difference (P < 0.05, FC > 1.2) between BMT and CON, but their significance between BMT and AMT was marginal (P < 0.3; likelihood ratio test). Each IR gene is indicated by a large circle, where blue indicates DecIR and red IncIR. DEGs are indicated by a small circle, where blue indicates downregulation and red indicates upregulation. Intron ratio for IR and gene expression levels for DEGs. In the bar graph, asterisks indicate statistically significant differences (*p < 0.05, **p < 0.01, ***p < 0.001, NS: not significant). (A) The pathway involving *NDUFA5*, *FOXRED1*, *NDUFV2* and *ATP5MGL* (72) regulates mitochondrial function. (B) The *TP73* (73) - *FAS* (74) - *CAV1* (75, 76) - *PDGFRA* (77) pathway is involved in inflammatory signaling. (C) The *CAV1* (75, 76) - *PLD2* (78) - *NAPEPLD* (79, 80) signaling pathway is involved in the regulation of lipid metabolism involving caveolae as a vital plasma-membrane sensor. (D) The *DDX5* (81, 82) - *ZWINT* (83) - *UBE2T* (84) pathway is involved in the amplification by ubiquitination of an inflammatory signal taken up by *DDX5* via the immune infiltration stimulated by *ZWINT*. (E) The *CERT1* (85) - *COL4A3* (86) - *ITGA9* (87) pathway is involved in anti-inflammatory responses. (F) The *HOXA1* (88) - *ROBO3* (89) - *NTN1* (90) signaling pathway regulates inflammation. (G) See text. (H) The *SMARCD2* (91, 92) - *BRD9* (93, 94) signaling pathway mediates inflammatory input activated by *BRD9*, thereby mediating granulopoiesis as an output through activation of *SMARCD2*. (I) See text. (J) The *EOGT* (95) - *DOCK6* (96) pathway regulates hematopoiesis. (K) In the main network, each position of the pathways (A–H) is indicated by a red outline.

## Discussion

The upregulated DEGs identified for depressed patients in this study were significantly enriched in GO terms associated with innate immunity, including phagocytosis. Approximately 50% of these genes encode immunoglobulin heavy chains (Figure 1B, Supplementary Table 1). In contrast, the downregulated DEGs were enriched in GO terms associated with adaptive immunity, including antigen presentation and T-cell function. This trend in depressed patients has been reported previously (3, 4).

### Intron fine-tuning model links IR to protein homeostasis

An important aspect of our study is that the data show that IR can be used as an alternative clinical means of diagnosing depression, in place of traditional DEG-based methods. Roughly speaking, GO terms enriched in IncIR genes corresponded to GO terms of upregulated DEGs, whereas GO terms enriched in DecIR genes corresponded to GO terms of downregulated DEGs. This correspondence can be better understood in the light of our recently proposed intron fine-tuning model (29). That is, an increase in IR leads to a decrease in the amount of mature cytoplasmic mRNA (and thus a decrease in cytoplasmic protein), whereas a decrease in IR leads to an increase in mature cytoplasmic mRNA (and thus an increase in cytoplasmic protein). In other words, IR is a function that optimizes the amount of cytoplasmic protein by detecting changes in the amount of protein required in the cytoplasm due to physiological adaptations that occur in response to stress. Namely, this is the molecular mechanism by which intracellular protein homeostasis is regulated.

### The sensor role of IR is evolutionarily conserved

As can be easily imagined from the model described above, genes that undergo IR have a sensor role. Indeed, in the literature describing the gene functions of many of the depression-related IR genes we identified, the article titles often include the word ‘sensor’ or ‘regulate’ (Table 1). Therefore, if an IR is observed in depressed patients for a gene whose sensor role is not reported in the literature, analysis of the IR gene may reveal a new, as yet unidentified, regulatory sensor role for the gene in clinically depressed patients.

In many cases, the IR genes we identified did not correspond to genes in the DEGs themselves. Of the 30 DEGs shown in Figure 1B, only one gene, *HLA-DRB1* (50), is actually subject to IR. DEGs are often reflect quantitative aspects, whereas IR genes are more qualitative. By analogy, the DEG is the soldier, the manual worker, whereas the IR genes are the commander in chief. A typical example of the qualitative difference between DEGs and IR genes is secreted proteins, which are sometimes detected as DEGs (indeed, half of the upregulated genes we identified were immunoglobulins; Supplementary Figure 1) but not as IR genes. This is because these proteins are secreted from cells via the Golgi and are therefore not captured by the homeostasis detection mechanism in the cytoplasm (29).

Based on our previous publications (27, 28, 29), it is likely that some genes (about 10%) of the 20,000-30,000 genes in the mammalian genome are originally ASSIGNED to play a sensor role by retaining introns during stress. In budding yeast, for example, introns may play a mediator role in monitoring the physiological state of the cell (51, 52). Thus, the sensor function of IR genes is likely to be evolutionarily conserved, as described in the present study. However, the mechanisms by which IR genes sense the physiological stresses experienced by cells and contribute to homeostasis (29) remain to be elucidated.

### IR is an excellent marker for diagnosing depressive states and is superior to DEG

Many researchers have analyzed DEGs between cases and controls to identify markers of depression. It has been found that even when the top 10 genes with the highest expression variability are examined, the data differ from experiment to experiment and do not agree (53). As our current study shows, IR variation is likely to be a more sensitive marker than DEGs for the diagnosis of depression. Consider the following practical interpretation: a 10% variation in immunoglobulin levels detected by DEG does not necessarily represent the physiological state of a particular individual, but a 10% decrease in the IR of the inflammation sensor STAT1 (39) could result in a 10% increase in the amount of cytosolic STAT1 protein, thus having a significant impact on immune homeostasis. The accumulation of various studies to date suggests that the cause of depression is polygenic as an explanation for the fact that DEG data fluctuate and do not agree from experiment to experiment (54), but the failure to find a better marker gene in the DEG is not so much that depression is polygenic, likely due to the nature of the DEGs themselves.

### Examples of markers for the diagnosis of depressive states

So, among the genes that cause IR, are there any that are particularly likely to be common markers for depression? Depression is an immune disease. Therefore, among the IR genes we identified, the 34 genes identified as immune-related (Figure 3D) are likely to be good candidates. In addition, the expression of immune-related genes changes in response to depression, and the IR genes most likely to interact with them would likely change with depression. Therefore, IR genes with a high ranking for interaction with immune-related genes, as shown in Figure 3FJK, also have great potential as markers for depression. Hub IR genes with many connections in the interactome (Figure 4B), which we discussed earlier, are also good candidates.

In terms of common markers of depression, a particular highlight of the present study is the detection of a number of cilia-related genes (32 genes; Figure 3C) as IR genes. This may reflect the functional stress state of cilia as antennae in leukocyte cells, although the presence of cilia on leukocyte cells is controversial (55). It is known that when dendritic cells present antigens to T cells, these two types of cells form structures known as immunological synapses, in which the internal structure of the T cell resembles that of cilia (56, 57, 58). Therefore, the observation of IR in ciliary genes may represent a failure of T cells to recognize antigens during depression. This interpretation is consistent with the observations that several T cell activation genes were downregulated (Figure 1E) and that those involved in the T cell signaling pathway were characterized as DecIR genes (Figure 2D).

Of the 1117 cilia genes currently known, IR was observed in 32 genes in this study (Figure 3C). Surprisingly, of the 37 cilia-specific genes currently known to cause Joubert syndrome, six (AHI1 (59), CELSR2 (60), CEP104 (61), IFT172 (62), NPHP1 (63), TMEM107 (64)) were found to be among the IR genes. In addition, IR was restored for four of these six genes in response to treatment with HKT (Figure 5AB). Thus, the Joubert syndrome genes appear to be frequently IR and highly responsive to HKT, suggesting that these genes are the best candidates for marker genes for depression. AHI1 and NPHP1 were also found to be involved in pathways for recovery, as described below (see Figures 5 and 6). Future clinical studies are awaited to confirm these possibilities, where it will be necessary to determine more reads during sequencing, assuming rMATS is used (rMATS requires approximately five times more sequencing reads than DEGs).

### Ten characteristic pathways represented by efficacies of HKT

Mapping of HKT-responsive IR genes and DEGs onto the IR-DEG interactome revealed 10 pathways for which IR of certain genes was recovered in concert with that of other genes. Although each of these 10 pathways deserves detailed investigation (most of them were newly discovered in this study), it is important to emphasize that, in each of these 10 pathways, changes in IR of one gene were linked to changes in IR or DEGs, forming a single functional unit (which we call a gear). We will discuss a few of these. The first is the *NPHP1*-*AHI1* pathway (Figure 6G). As mentioned above, these two genes are involved in cilia function, and mutations cause a ciliopathy called Joubert syndrome (59, 65, 66, 67). A yeast two-hybrid analysis revealed that jouberin (encoded by *AHI1*) can interact with nephrocystin (encoded by *NPHP1*) (68). The two proteins form a heterodimer, and a mutation in *AHI1* (V443D) that prevents heterodimer formation alters the intracellular localization of AHI1 and NHPH1 so that the two proteins can, although not always, behave as if they were one protein (59). It is interesting to note that the IR of these two mRNAs is reduced in depression and that IR of both mRNAs was returned to that of the healthy state in response to treatment with HKT (Figure 6G). In other words, the mRNAs transcribed from these two genes seem to be under the same control mechanism of RNA processing, as if they were the same mRNA. These observations remind us of a model we recently proposed, i.e., a novel mechanism may exist that senses the correct level of functional proteins in the cytoplasm and transmits this information to the nucleus to regulate the level of IR (29). If such a mechanism exists, it would mean that IR in of *AHI1* and *NPHP1* is regulated by a common factor.

In the case of the *MYLK*-*MYH10* pathway (Figure 6I), inflammatory inputs activate MYLK and phosphorylate the L-chain of myosin (47). This causes the contracted L-chain of myosin to transmit information to the H-chain, which in turn regulates the copy number of mitochondrial DNA, which is tightly bound to the non-muscle H-chain (45). In this biological GEAR, the input is inflammation and the output is the control of the number of copies of mitochondrial DNA. The GEAR function in these ten pathways, including the two already postulated, need to be demonstrated biochemically, but brief outlines of the hypothetical pathways are given in the legend of Figure 6.

Figure 6K shows the mapping of the 10 pathways for which IR was restored by HKT on the IR-DEG interactome described in this study. This interactome was generated using data from depressed patients and controls only. Thus, if a drug other than HKT (i.e., with a different effect) was used, new pathways restored by the drug could be detected. Accordingly, this IR-DEG interactome should be useful for assessing the efficacy of individual drugs, including herbal medicines, and for identifying new pathways affected by drugs.

In summary, our results show that IR can be an excellent marker of depression. The combination of network analysis and analysis of drug-responsive IR genes may also reveal new pathways of drug action. The strategy presented here is not limited to the analysis of depression, but rather could be applied to any disease.

## Data Availability

All data produced in the present study are available upon reasonable request to the authors

## Authors’ Contributions

Norihiro Okada conceived, supervised and validated the IR project and wrote and finalized the manuscript. Kenshiro Oshima analyzed and visualized the data and wrote the Materials and Methods and legends. Akiko Maruko analyzed the PCR data. Mariko Sekine suggested the application of the IR method to human subjects. Naoki Ito isolated PBMCs. Akino Wakasugi analyzed the human data. Eiko Mori and Hiroshi Odaguchi designed and performed the clinical research. Yoshinori Kobayashi organized the human COI project to investigate the effectiveness of herbal medicine. All authors read, revised, edited, and approved the final manuscript.

