## Supplementary Figure 2 for "Intron retention as an excellent marker for diagnosing depression and for discovering new potential pathways for drug intervention" (from another version of this preprint: version 2, 2024-04-02)

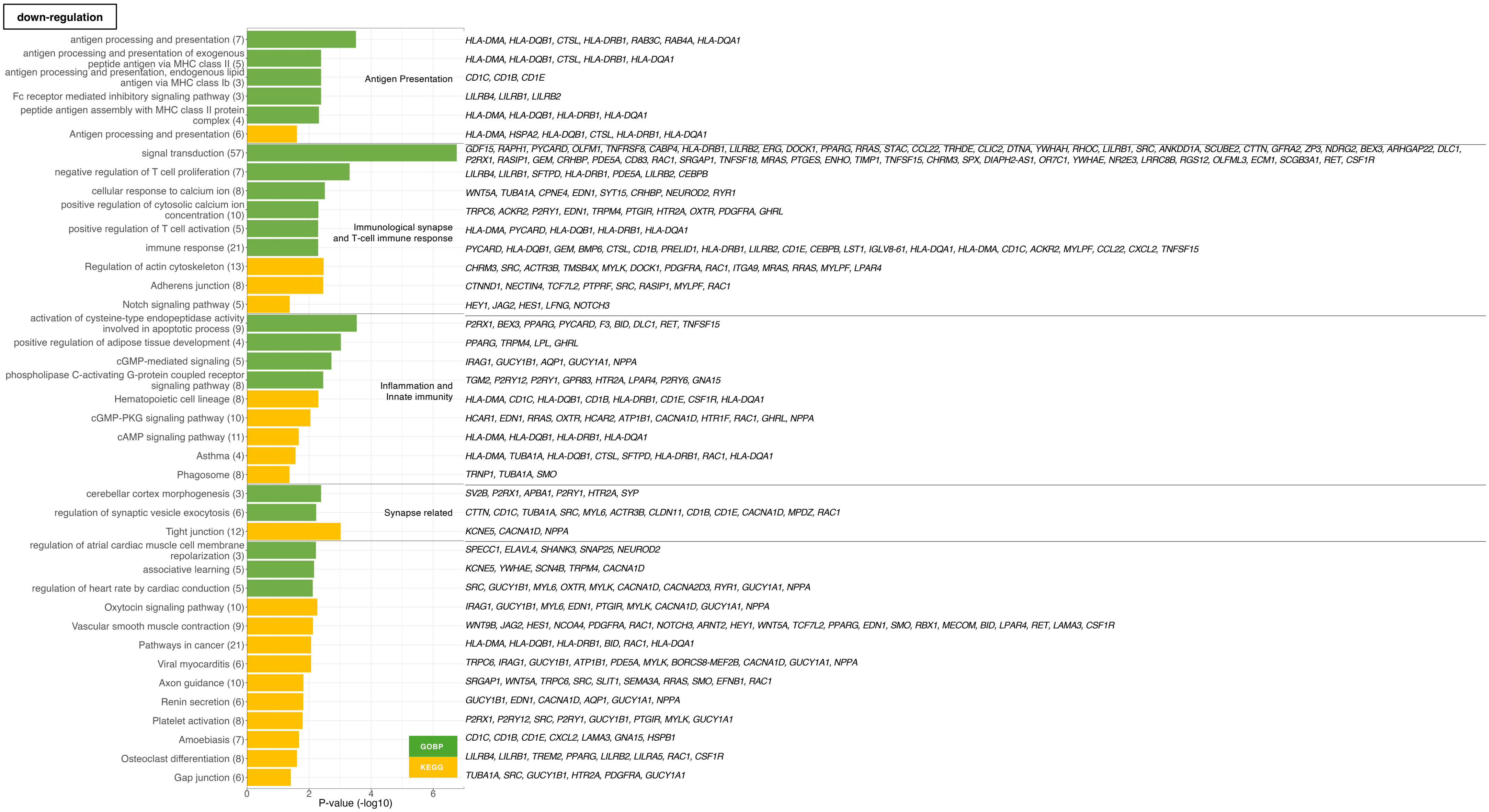


**Supplementary Figure 2. Bar chart of enrichment analysis of biological process gene ontology and KEGG pathway terms using down-regulated genes in BMT.**
